# Dengue spatiotemporal patterns in Minas Gerais, Brazil, 2014 – 2023: regional epidemic forces dominate over the environmental impact of the Brumadinho dam collapse

**DOI:** 10.64898/2026.05.19.26353615

**Authors:** Gabriel da Rocha Fernandes, Aline Bruna Martins Vaz, Paula Luize Camargos Fonseca, Wanderson Kleber Oliveira, Eric Roberto Guimarães Rocha Aguiar, Bruna Coelho Lopes, César Rossas Mota-Filho, Múcio Leão Pessoa Castro, Carlos Ernesto Starling

**Affiliations:** Instituto René Rachou - Fiocruz Minas Belo Horizonte BR; Universidade Professor Edson Antônio Velano (UNIFENAS) Department of Medical Educaiton Belo Horizonte BR; Universidade Federal de Minas Gerais (UFMG) Department of Genetics, Ecology and Evolution Belo Horizonte BR; Hospital das Forças Armadas Brasilia BR Centro Universitário (Uniceplac) Medical School Brasília BR; Universidade Estadual de Santa Cruz (UESC) Department of Engineering and Computation Ilhéus BR; Universidade Federal de Minas Gerais (UFMG) Department of Sanitary and Environmental Engineering Belo Horizonte BR; Pathos Ltda. Belo Horizonte BR; Infection Control Ltda. Belo Horizonte BR

## Abstract

**Background:** Dengue is a major public health problem in Brazil, and Minas Gerais is one of the states with the highest burden. In January 2019, the Brumadinho dam collapse released about 12 million cubic meters of iron ore tailings into the Paraopeba River basin, causing environmental disturbance that could plausibly affect vector habitats and dengue transmission. We evaluated the spatiotemporal dynamics of dengue in Minas Gerais from 2014 to 2023 and tested whether the disaster was associated with changes in affected municipalities.

**Methods:** We performed an ecological spatiotemporal analysis using dengue notifications from SINAN for all municipalities in Minas Gerais (2014–2023). Municipalities were classified as Paraopeba basin, regional controls, or state controls. Temporal similarity was assessed using Pearson correlation-based hierarchical clustering and non-metric multidimensional scaling (NMDS). Sources of variation were examined with PERMANOVA and principal component analysis (PCA). A linear mixed-effects model with municipality as a random effect was used to test changes after 2019, with pre/post contrasts estimated from marginal means.

**Results:** Dengue showed strong temporal synchrony across the state, with major epidemic peaks in 2015–2016, 2019, and 2023. Health region explained 31.5% of the variation in temporal incidence profiles (p = 0.001), whereas Paraopeba basin status explained no significant variation (p = 0.998). No temporal cluster was enriched for municipalities in the Paraopeba basin. PCA identified 2023, 2019, and 2016 as the main years driving variability. In the mixed model, year was significant (p < 0.001), but Paraopeba basin status and its interaction with time were not. Incidence increased significantly after 2019 in non-exposed municipalities (p < 0.001), but not in basin municipalities (p = 0.088).

**Conclusions:** Dengue dynamics in Minas Gerais were driven mainly by regional and state-wide epidemic processes, with no significant independent effect of the Brumadinho dam collapse on notified dengue patterns.

## 1. Introduction

Dengue, caused by dengue virus (DENV), is the most worrying mosquito-borne viral disease worldwide. Transmitted primarily by *Aedes aegypti* across tropical and subtropical regions (Churakov et al. 2019; Bhatt et al. 2013; Lim et al. 2025). Brazil bears the heaviest dengue burden globally, accounting for the largest share of reported cases and a substantial proportion of infections across the Americas (San Martín et al. 2010). Over recent decades, the country has experienced increasingly frequent and intense epidemics, shorter inter-epidemic intervals, and a state of hyperendemicity characterized by the co-circulation of all four DENV serotypes (Andrioli, Busato, and Lutinski 2020). More recently, the resurgence of DENV-3 in 2023 underscored the persistence of epidemic risk even in populations with prior serotype exposure (Adelino et al. 2024).

The transmission dynamics of dengue are inherently complex and spatially heterogeneous, shaped by the interaction of climatic, environmental, demographic, and socioeconomic factors (Teurlai et al. 2015). These drivers produce distinct regional transmission profiles and give rise to large-scale spatiotemporal patterns, such as seasonal “travelling waves” of dengue spread across Brazil (Bhatt et al. 2013; Churakov et al. 2019). Previous studies have further demonstrated that dengue transmission in Brazil follows structured spatiotemporal patterns with strong geographic clustering and directional spread across regions, reinforcing the importance of spatial connectivity and regional interactions in shaping epidemic dynamics (Lowe et al. 2021). Consequently, spatiotemporal approaches have become essential tools for understanding dengue epidemiology, enabling the identification of high-risk areas and the characterization of underlying transmission structures across diverse geographic contexts.

In this context, the state of Minas Gerais, located in southeastern Brazil, represents a relevant setting for investigating dengue dynamics. The state consistently reports a high burden of cases, with transmission patterns strongly associated with urbanization and social vulnerability (Andrioli, Busato, and Lutinski 2020; Bohm et al. 2024). In January 2019, the Brumadinho dam collapse released approximately 12 million cubic meters of mining tailings into the Paraopeba River basin, causing substantial environmental disturbance (Vergilio et al. 2020). The resulting mudflow disrupted riparian landscapes, affected water supply systems, and may have altered aquatic breeding habitats for *Aedes aegypti*, creating conditions that could plausibly modify local dengue transmission dynamics. Whether these environmental changes produced a detectable epidemiological signal remains, however, an open empirical question.

Concurrent large-scale disruptions complicate this picture further. The COVID-19 pandemic led to reductions in dengue vector control activities and contributed to case underreporting across Brazil (Rotondo De Araújo et al. 2023; Borre et al. 2022; Oliveira Roster et al. 2024). However, these effects are considered secondary in the present study, which focuses primarily on environmental disruption as a potential driver of epidemiological change. Importantly, structural limitations of passive surveillance systems, including reduced sensitivity in detecting dengue cases, further complicate the interpretation of temporal trends (Coelho et al. 2016).

Given these overlapping environmental and systemic factors, there is a need for robust spatiotemporal analyses that integrate epidemiological data with geographic and contextual variables. In this study, we evaluate the spatiotemporal distribution of dengue in Minas Gerais from 2014 to 2023, testing the hypothesis that the 2019 dam collapse may have influenced the epidemiological dynamics of arboviral transmission in affected municipalities, while considering broader regional patterns and temporal variability.

## 2. Methods and Materials

### 2.1. Study design and data sources

We conducted an ecological spatiotemporal analysis of dengue incidence in Minas Gerais, Brazil, from 2014 to 2023. Case notification data were obtained from the Brazilian Notifiable Diseases Information System (SINAN), including date of symptom onset and municipality of residence. Records were aggregated at the municipality-year level and linked to demographic data using IBGE municipality codes .

The study period was divided into pre-event (2014-2018) and post-event (2019-2023) intervals relative to the Brumadinho dam collapse of 25 January 2019. All 67 municipalities included in the study were selected based on their involvement with the dam collapse - either directly or indirectly - and classified into two groups for the impact analysis. The “Paraopeba basin” group (n = 26) comprises municipalities through which the Paraopeba River flows and that were formally designated as affected in the post-disaster governance process; this designation arose from a technical-political agreement under which these municipalities received reparation investments to address the environmental and socioeconomic consequences of the collapse. The “regional controls” group (n = 41) comprises municipalities that belong to the same administrative health regions as the Paraopeba basin municipalities but were not included in the affected designation. This design allows comparison of temporal dengue dynamics within shared regional contexts, controlling for health-region-level determinants of transmission. A third group - state-level controls (all remaining Minas Gerais municipalities) - was included in the descriptive analyses to contextualise incidence trends at the state level but was not used in the formal impact analysis. The municipality-level characteristics of all study groups are summarised in Supplementary Table S1.

### 2.2. Incidence and standardization

Annual dengue cases were computed for each municipality, with missing values set to zero. Incidence rates (per 1,000 inhabitants) were calculated using 2022 census population data. To enable comparison of temporal patterns independently of absolute magnitude, incidence was standardized using municipality-specific Z-scores.

### 2.3. Pattern recognition and clustering

Temporal similarity between municipalities was assessed using a distance matrix defined as 1 minus Pearson correlation. Hierarchical clustering was applied, and the optimal number of clusters was determined using the silhouette method (Rousseeuw 1987) . Cluster composition was evaluated using Fisher’s exact test with Benjamini–Hochberg correction for multiple comparisons (Benjamini and Hochberg 1995).

### 2.4. Multivariate analysis

Differences in epidemiological profiles were quantified using permutational multivariate analysis of variance (PERMANOVA) based on the distance matrix (Anderson 2001), including Paraopeba basin status and health region as explanatory variables. Ordination was visualized using non-metric multidimensional scaling (NMDS) (Kruskal 1964). Principal component analysis (PCA) was applied to identify the years contributing most to variability in incidence patterns (Jolliffe 2014).

### 2.5. Statistical modeling

To test whether epidemiological patterns changed after 2019, a linear mixed-effects model was fitted with municipality as a random effect (Bates et al. 2015). The outcome was the Z-score of incidence, with year (categorical), Paraopeba basin status, health region, and their interactions as fixed effects. Model significance was assessed using Type III ANOVA, and contrasts between pre- and post-2019 periods were estimated using marginal means (*emmeans*) to evaluate within- and between-group differences.

### 2.6. Software

All analyses were performed in R (R Core Team, 2023), using the *vegan* (Oksanen et al. 2001), *lme4* (Bates et al. 2015), and *emmeans* packages (Lenth and Piaskowski 2017).

### 2.7. Study size

No formal a priori sample size calculation was performed. The study population was determined by the scope of the dam disaster: all 67 municipalities involved with the Brumadinho dam collapse — either directly as Paraopeba basin municipalities or indirectly as regional controls — were included in their entirety. This constitutes a census of the municipalities relevant to the exposure rather than a sample, making conventional power calculations inapplicable. The study period spans 2014 to 2023, the longest interval currently available from the SINAN surveillance system (which began systematic nationwide electronic recording in 2014), providing five years of pre-event data (2014-2018) and five years of post-event data (2019-2023).

## 3. Results

### 3.1. Temporal patterns of dengue incidence

Across all municipalities, dengue incidence exhibited marked temporal variability, with epidemic peaks occurring primarily in 2015–2016, 2019, and 2023 (Supplementary Fig. S1). These peaks were consistently observed across municipalities regardless of exposure status, indicating strong state-wide temporal structuring of dengue dynamics.

**Supplementary Fig. S1.**
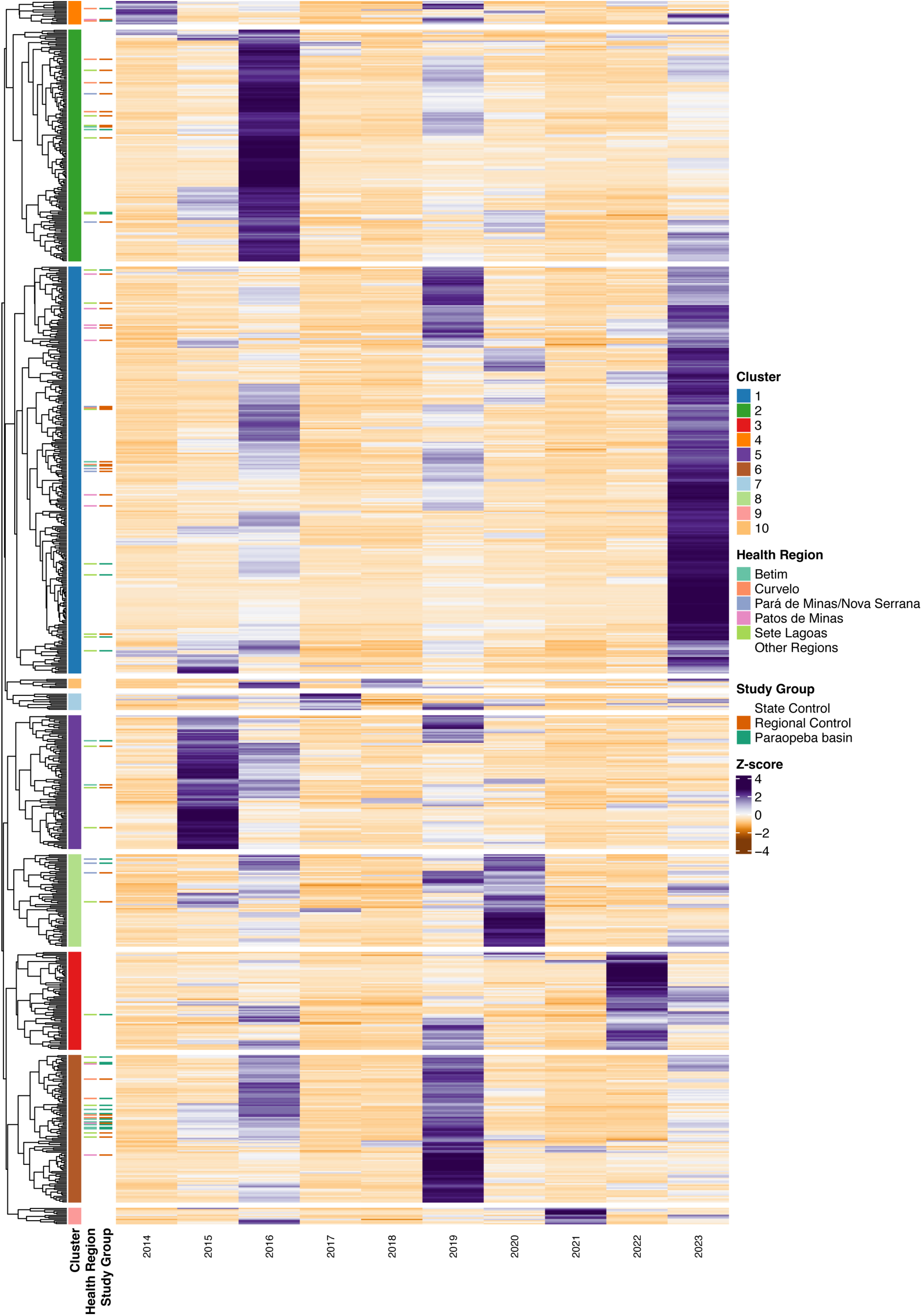
Heatmap of standardized dengue incidence (Z-scores) across all municipalities of Minas Gerais from 2014 to 2023. Each row represents one municipality and each column represents one year. Purple shading indicates incidence above the municipality-specific historical mean; orange indicates below-average incidence. Epidemic peaks in 2015–2016, 2019, and 2023 are visible as horizontal bands of purple spanning most municipalities simultaneously.

Monthly analysis further revealed that municipalities within the same health region exhibit highly synchronized temporal patterns, which largely overlap with those observed in other regions of the state (Fig. 1). Epidemic peaks occur simultaneously across regions, reinforcing the presence of shared large-scale temporal drivers of dengue transmission rather than localized dynamics.

**Fig. 1.**
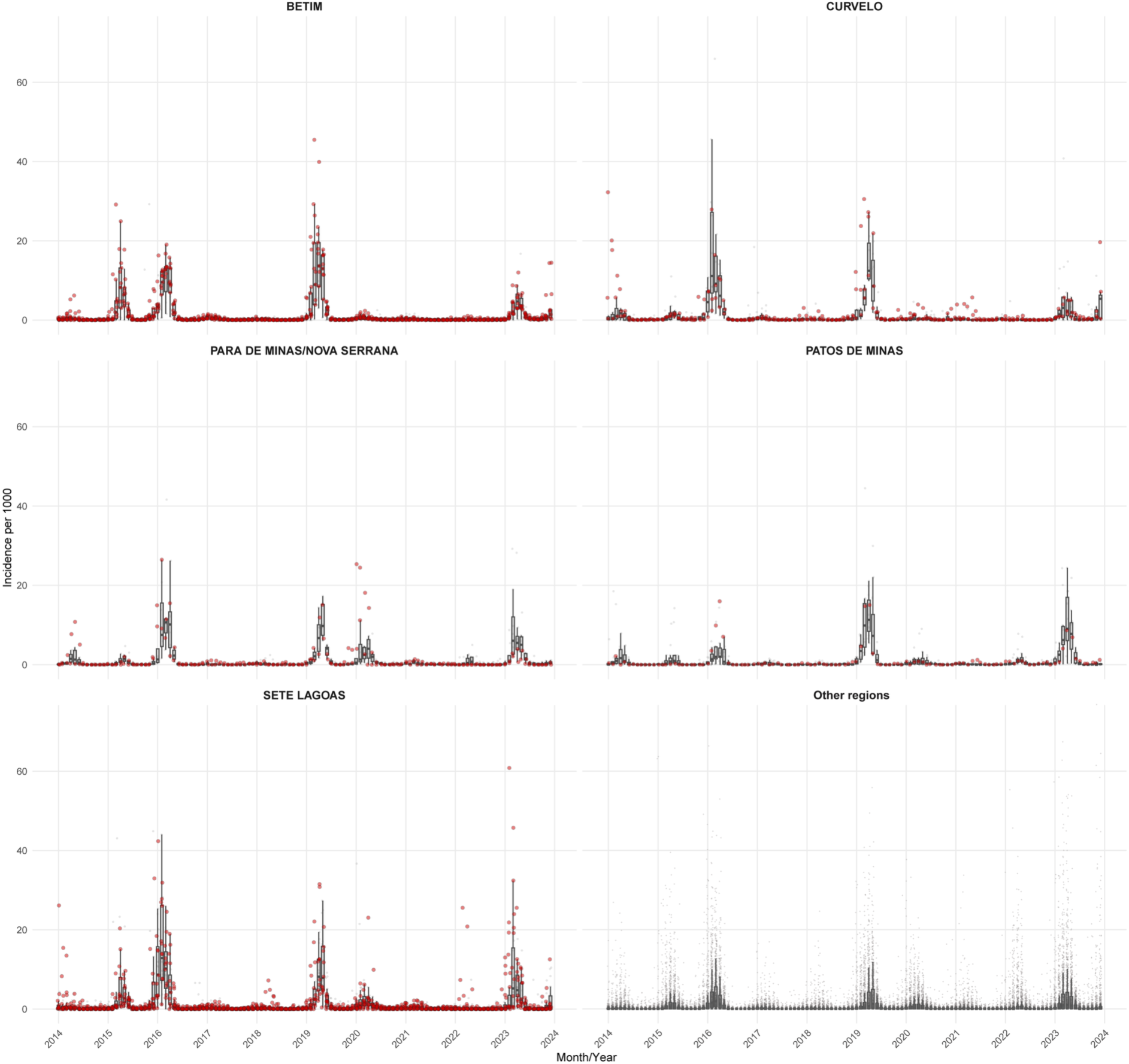
Monthly dengue incidence across health regions of Minas Gerais, 2014–2023. Each panel corresponds to one health region. Incidence is expressed per 1,000 inhabitants. Red points indicate municipalities within the Paraopeba River basin (exposed group); grey points indicate control municipalities (regional controls within the same health region, or state-level controls in the “Other regions” panel).

Comparison of aggregated case counts before and after 2019 showed a broad increase in dengue incidence across Minas Gerais and Brazil (Supplementary Fig. S2), consistent with the large national outbreaks that characterized the post-event period, particularly in 2019 and 2023.

**Supplementary Fig. S2.**
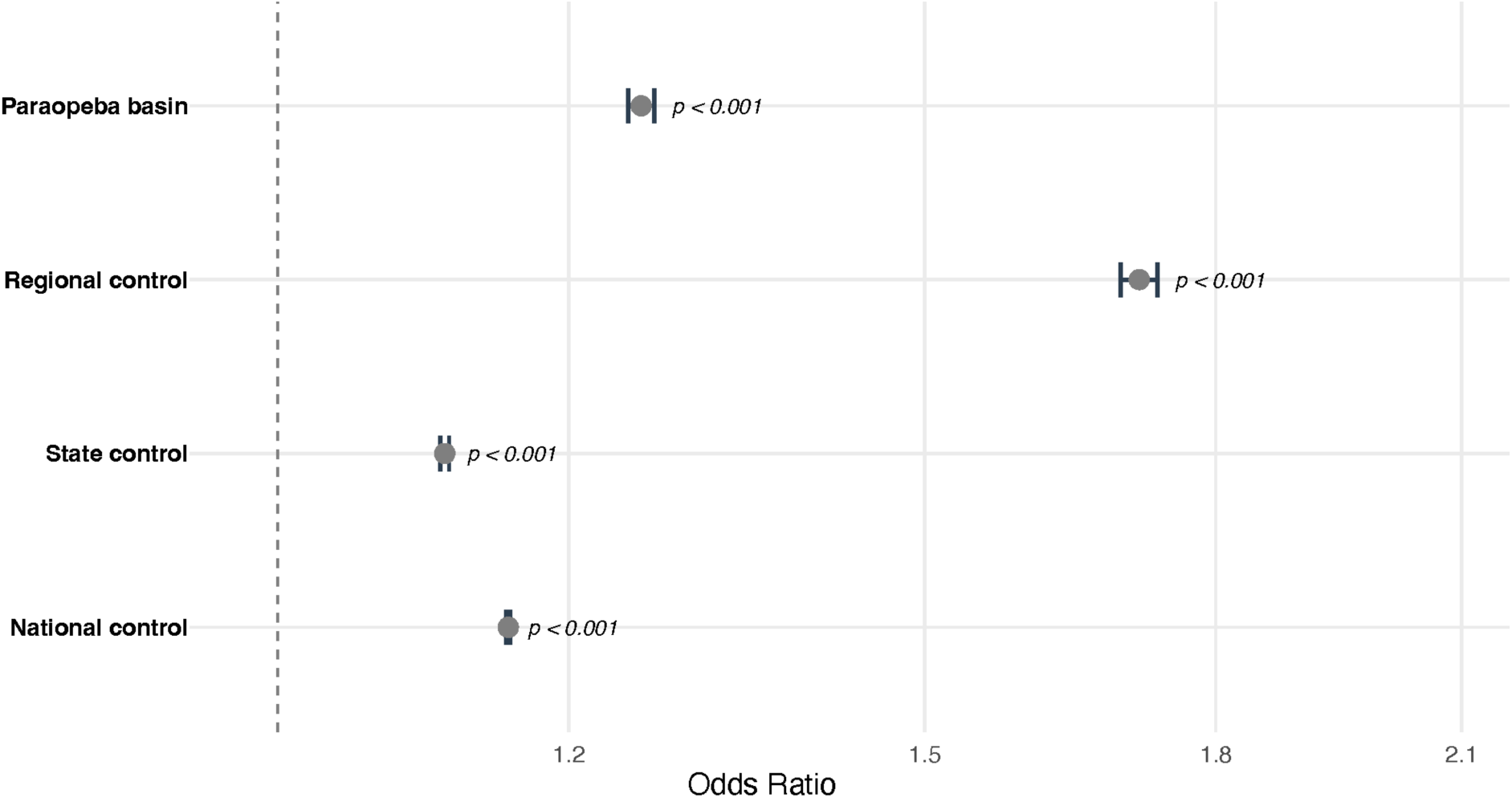
Forest plot comparing mean dengue incidence rates (per 1,000 inhabitants) before (2014–2018) and after (2019–2023) the dam collapse across four study groups: Paraopeba basin municipalities, regional controls, state-level controls, and national controls. Points represent group means and error bars represent 95% confidence intervals.

### 3.2. Similarity structure and clustering of epidemiological profiles

Hierarchical clustering of temporal incidence profiles identified five distinct municipality groups across the combined sample of Paraopeba basin and control municipalities. The distance matrix showed clear within-cluster similarity (Supplementary Fig. S3), and Z-score heatmaps confirmed that municipalities within the same cluster shared synchronized epidemic peaks and troughs (Fig. 2).

**Fig. 2.**
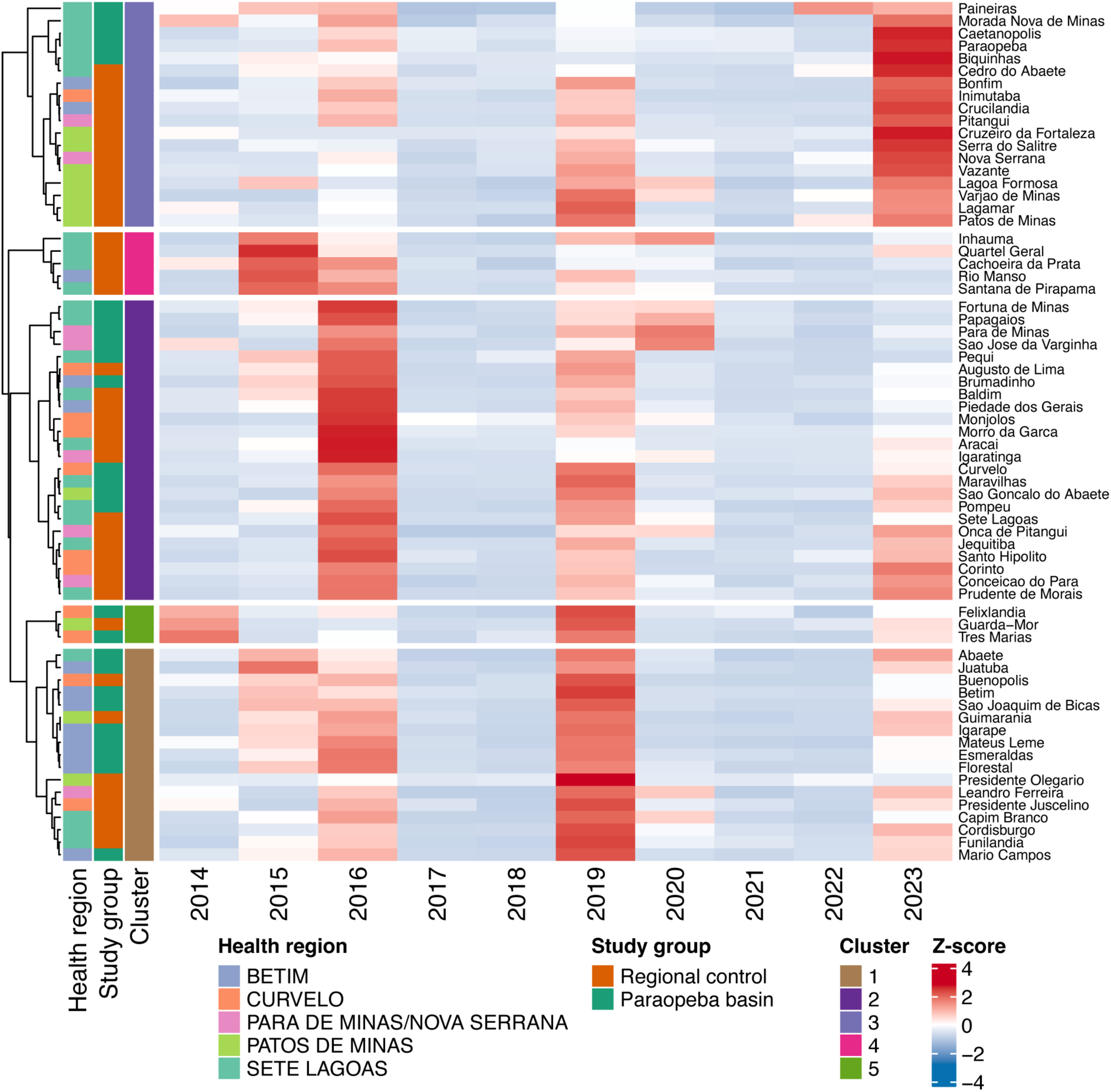
**Clustered heatmap of standardized dengue incidence (Z-scores) for Paraopeba basin and control municipalities. Rows represent municipalities grouped into five clusters by hierarchical clustering (cluster membership indicated by the colour bar on the left); columns represent years (2014–2023). Red shading indicates above-average incidence and blue indicates below-average incidence relative to each municipality’s historical mean.**

**Supplementary Fig. S3.**
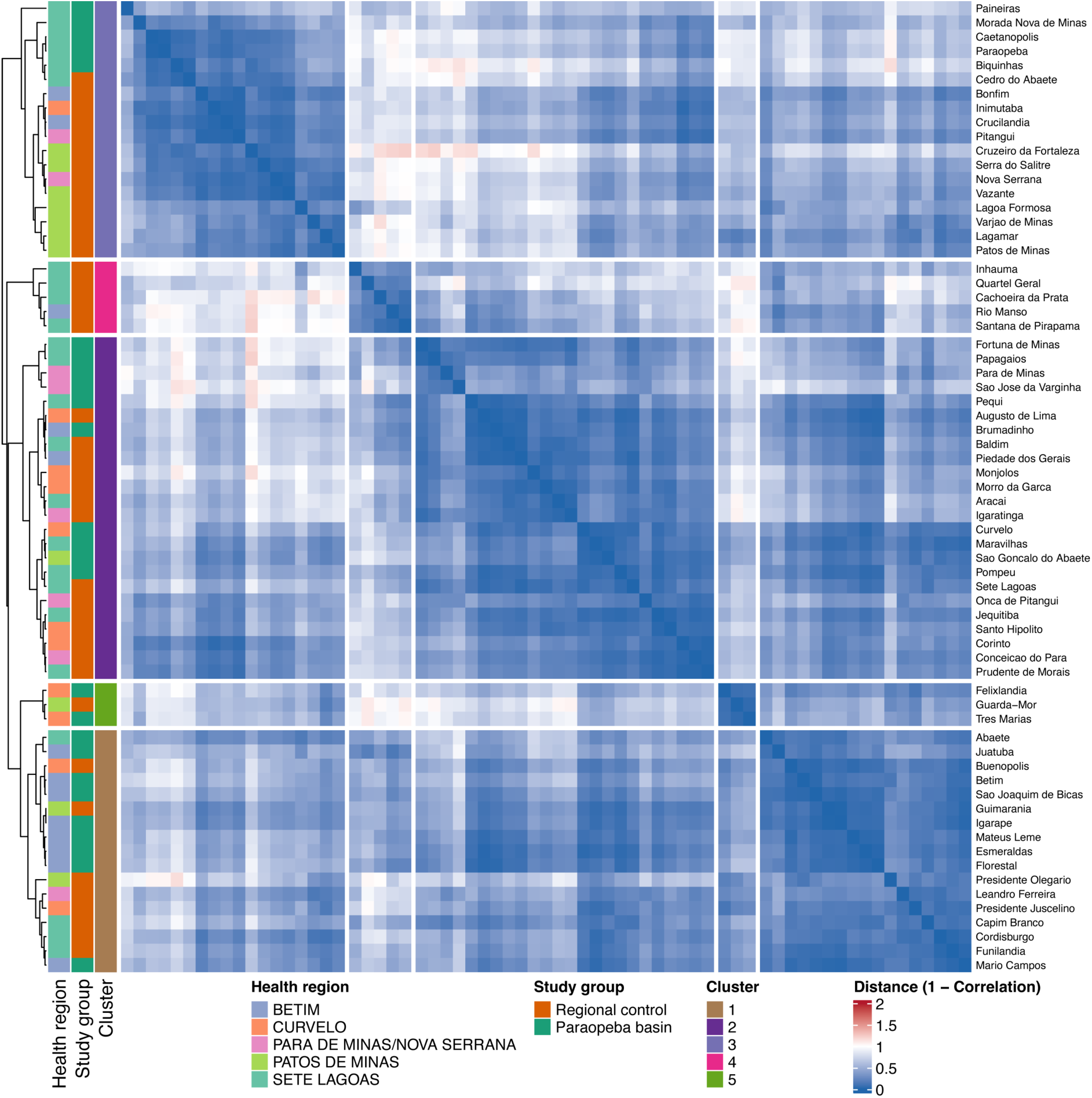
Heatmap of the pairwise distance matrix (1 − Pearson correlation) between municipalities, ordered by hierarchical clustering. Darker colours indicate greater dissimilarity; lighter colours indicate greater temporal similarity. The five cluster boundaries identified by the silhouette method are visible as blocks along the diagonal.

**Municipalities within the same cluster share similar interannual incidence trajectories.**

Cluster enrichment analysis revealed no significant overrepresentation of Paraopeba basin municipalities in any cluster after multiple-testing correction. In contrast, two clusters showed suggestive health-region enrichment (q < 0.1): cluster 1 was enriched for the Betim region and cluster 3 for the Patos de Minas region, indicating that regional structure contributes meaningfully to the similarity of temporal incidence profiles (Supplementary Table 2).

### 3.3. Multivariate structure of dengue dynamics

PERMANOVA analysis demonstrated that Paraopeba basin status did not significantly explain variability in temporal incidence profiles (R² ≈ 0%; p = 0.998), whereas health region accounted for a substantial proportion of the observed variation (R² ≈ 31.5%; p = 0.001).

Ordination using NMDS confirmed these findings, showing no clear separation between exposed and non-exposed municipalities, while municipalities tended to cluster according to health region (Fig. 3).

**Fig. 3.**
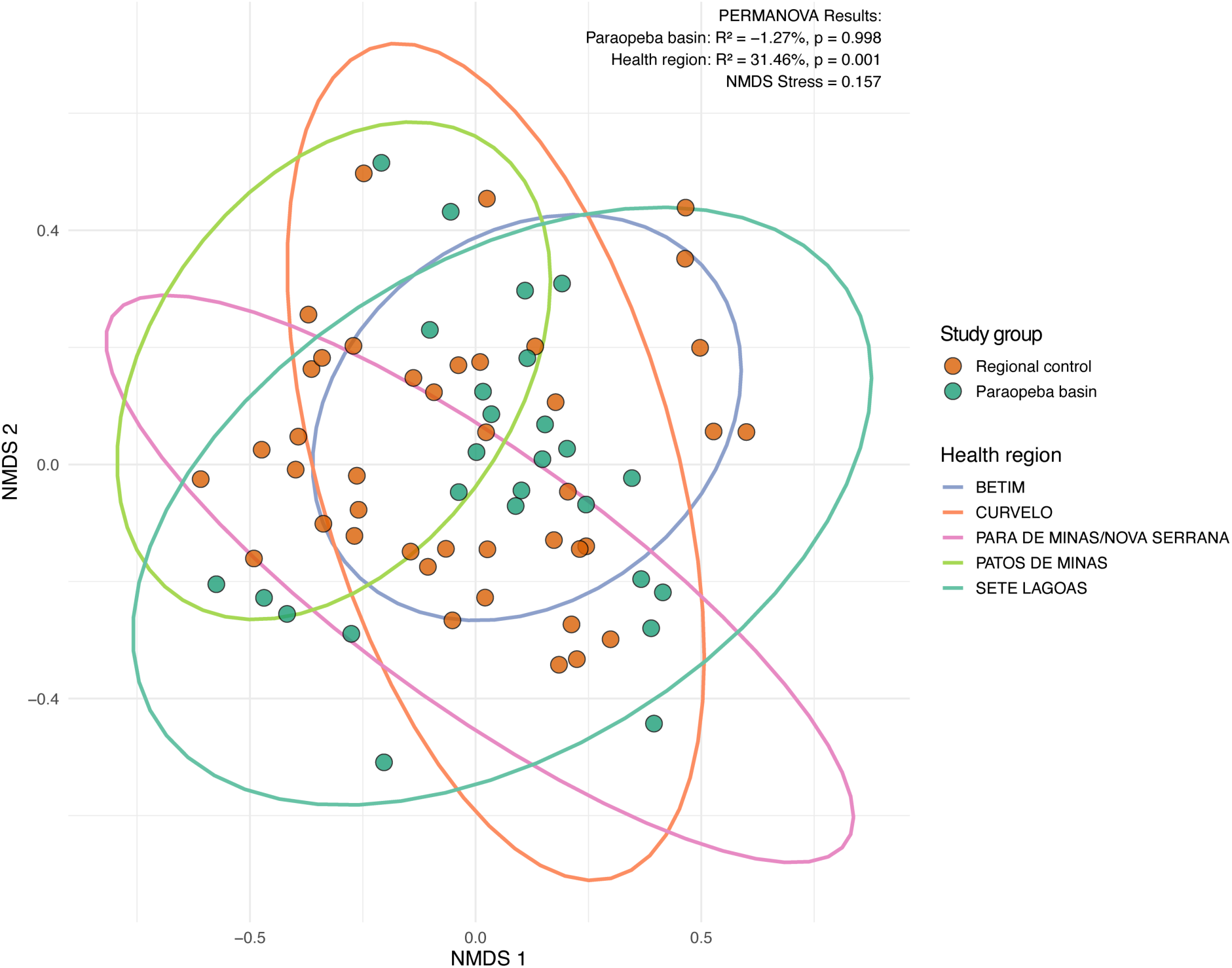
**Non-metric multidimensional scaling (NMDS) ordination of municipalities based on pairwise temporal incidence dissimilarity (1 − Pearson correlation). Each point represents one municipality. Point colour indicates Paraopeba basin status (exposed vs. non-exposed). Ellipses represent 95% confidence regions for each health region. Shorter distances between points reflect greater similarity in interannual incidence trajectories.**

Principal component analysis further indicated that interannual variability was driven primarily by epidemic years, with 2023, 2019, and 2016 contributing most strongly to the dispersion of municipalities in multivariate space (Fig. 4; Table 1) .

**Fig. 4.**
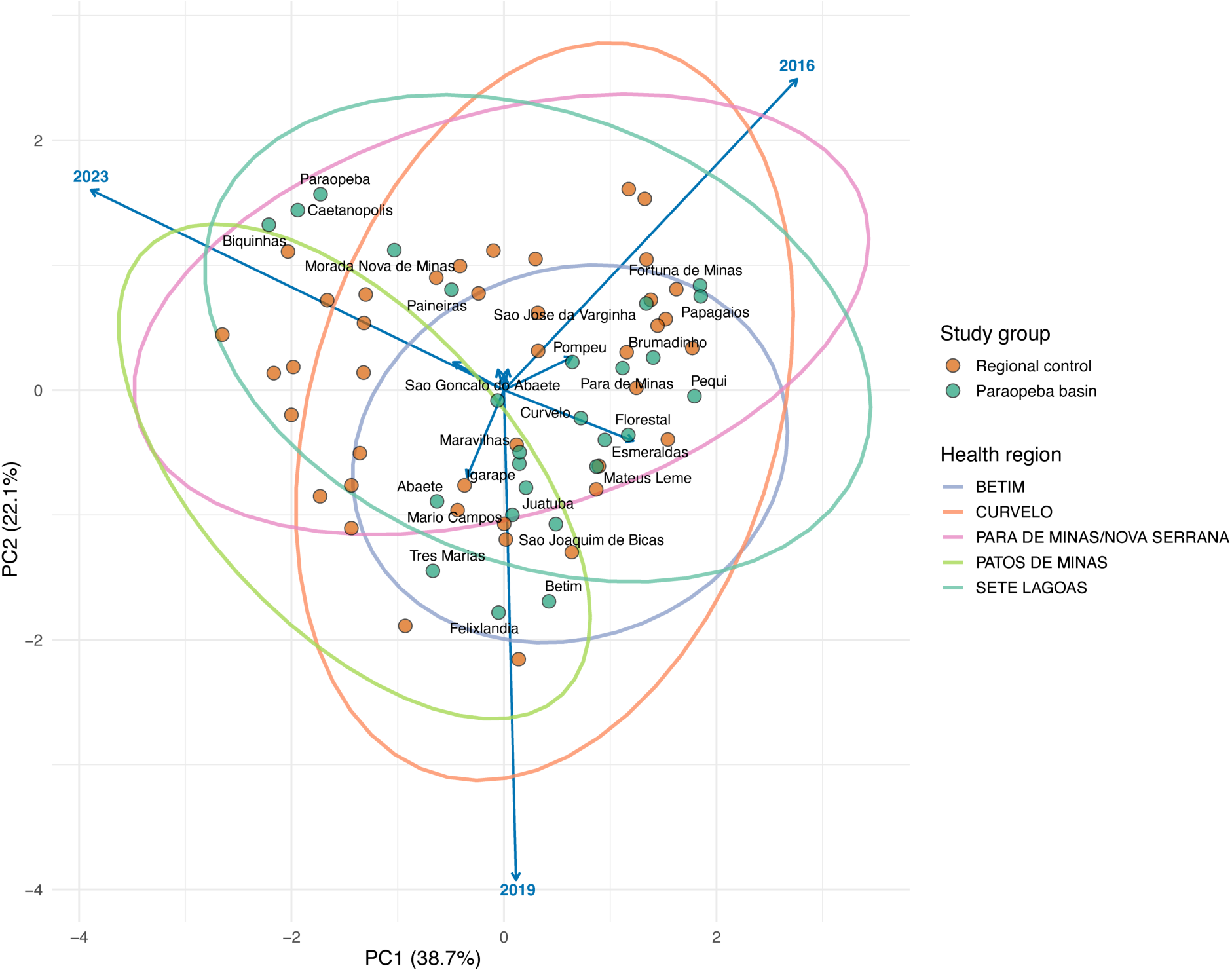
Principal component analysis (PCA) biplot of standardized dengue incidence profiles. Each point represents one municipality; points colours correspond to the study group, while elipses colours correspond to health region. Arrows represent years (2014–2023) as variables; arrow length is proportional to the contribution of that year to overall multivariate variability. The five years with the strongest loadings are listed in. **Table 1**.

**Table 1:** The 5 variables with the strongest vector loadings in the PCA.

| Year | Importance | PC1 | PC2 |
| --- | --- | --- | --- |
| 2023 | 0.841 | -0.778 | 0.321 |
| 2019 | 0.785 | 0.023 | -0.784 |
| 2016 | 0.744 | 0.552 | 0.498 |
| 2015 | 0.257 | 0.244 | -0.081 |
| 2014 | 0.156 | -0.069 | -0.139 |

### 3.4. Impact of the 2019 environmental event

The linear mixed-effects model confirmed that year was the dominant predictor of dengue incidence (p < 0.001), while Paraopeba basin status and its interaction with time were both non-significant (Table 2).

**Table 2:** Mixed model ANOVA (Type III)

|  | Chisq | Df | p-value |
| --- | --- | --- | --- |
| (Intercept) | 5.278 | 1 | 0.022 |
| Year (factor) | 70.674 | 9 | 0.000 |
| Paraopeba basin | 0.014 | 1 | 0.905 |
| Health region | 2.852 | 4 | 0.583 |
| Year:Paraopeba basin | 12.290 | 9 | 0.197 |
| Year:Health region | 135.129 | 36 | 0.000 |
| Paraopeba basin:Health region | 7.289 | 4 | 0.121 |
| Year:Paraopeba basin:Health region | 81.015 | 36 | 0.000 |

Contrasts comparing pre- and post-2019 periods showed a significant change in incidence patterns among non-exposed municipalities (p < 0.001), whereas no significant change was detected among municipalities within the Paraopeba basin (p = 0.088) (Table 3).

**Table 3:** Evaluation of pattern change (After vs. before) inside each study group.

| Contrast | Study group | Estimate | SE | DF | p-value |
| --- | --- | --- | --- | --- | --- |
| Before - After | Regional control | -0.245 | 0.060 | 513 | 0.000 |
| Before - After | Paraopeba basin | -0.172 | 0.101 | 513 | 0.088 |

These results indicate that the observed increase in dengue incidence after 2019 reflects a broader temporal phenomenon affecting the entire state, rather than a localized effect associated with environmental exposure in the Paraopeba basin. This pattern is further illustrated by the predicted temporal trends from the mixed-effects model (Fig. 5).

**Fig. 5:**
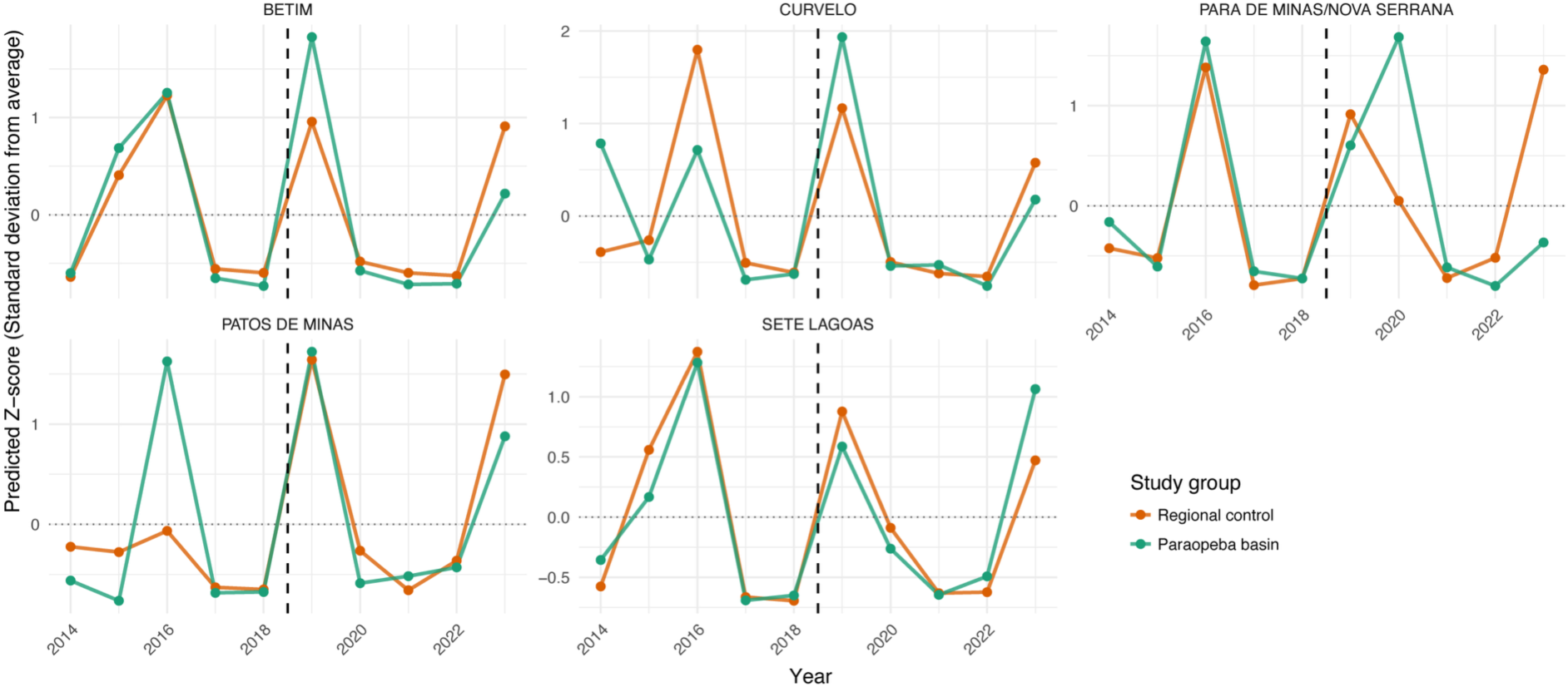
**Predicted temporal trends in standardized dengue incidence from the mixed-effects model. Lines represent estimated mean Z-scores for municipalities inside and outside the Paraopeba basin. The dashed vertical line indicates the year 2019.**

## 4. Discussion

This study evaluated the spatiotemporal dynamics of dengue incidence in Minas Gerais between 2014 and 2023, testing whether the 2019 Brumadinho dam collapse modified the epidemiological pattern of dengue transmission in municipalities within the Paraopeba River basin. The main finding was that, despite a significant overall increase in dengue incidence across the state in the post-event period, no distinct epidemiological signature attributable to the dam disaster was detected among exposed municipalities. Instead, health region emerged as the dominant structuring factor of dengue dynamics, and temporal variation was driven by state- and national-level epidemic forces rather than by local environmental disruption.

The predominance of health region as an explanatory factor for the clustering of epidemiological profiles is consistent with well-established evidence that dengue transmission in Brazil is organized by large-scale spatiotemporal processes, including travelling epidemic waves driven by climate seasonality and human mobility (Churakov et al. 2019). The strong regional structuring observed in our PERMANOVA analysis (R² ≈ 31.5%; p = 0.001) aligns with observations from other Brazilian states demonstrating that dengue dynamics follow mesoregional and health-administrative boundaries shaped by shared climate exposure, vector ecology, and population connectivity (Baes Pereira et al. 2025). This regional coherence suggests that the health region framework captures meaningful epidemiological units where shared determinants - including temperature, rainfall, urbanisation patterns, and healthcare access -modulate dengue risk homogeneously. The synchrony of epidemic peaks observed simultaneously across health regions of Minas Gerais, regardless of Paraopeba basin status, further supports this interpretation and corroborates findings from national-scale analyses of dengue spread in Brazil (Churakov et al. 2019; Lowe et al. 2021).

The three epidemic years that dominated multivariate variability in the PCA — 2023, 2019, and 2016 — correspond to known large-scale dengue outbreaks in Brazil and Minas Gerais. The 2023 epidemic was particularly severe: approximately 260,000 dengue cases were confirmed in Minas Gerais alone, a 3.5-fold rise relative to 2022 (De Jesus et al. 2024). This peak was associated with the co-circulation of multiple DENV lineages and the simultaneous re-emergence of chikungunya, suggesting complex arboviral dynamics driven by large-scale viral introductions and broad population susceptibility (De Jesus et al. 2024). The 2019 peak, coinciding with the year of the dam collapse, was equally a state-wide phenomenon not confined to the Paraopeba basin, as shown by the parallel temporal trajectories of exposed and non-exposed municipalities in the mixed-effects model. This temporal overlap between the disaster and a major national outbreak is a critical confounder: any apparent local increase in dengue in 2019 cannot be attributed to the dam collapse without accounting for the concurrent epidemic background.

These results differ, at least in part, from a recent ecological analysis of infectious disease notifications following the Mariana (2015) and Brumadinho (2019) dam disasters, which reported dengue increases of more than 100% in both affected municipalities (Machado et al. 2025). Those estimates, however, relied on simple pre-post comparisons without contemporaneous controls, making it difficult to disentangle any local disaster effect from the state-wide dengue surge that already characterized 2019. By controlling for health region effects and using a mixed-effects framework with an explicit exposed/unexposed contrast, our design is better positioned to isolate a disaster-specific signal. Neither the Year:Paraopeba basin interaction (p = 0.197) nor the pre-/post-2019 contrast for basin municipalities (p = 0.088) reached statistical significance, providing little support for the hypothesis that the dam collapse independently altered local dengue dynamics beyond the state-wide trend.

It is worth considering why a disaster of this magnitude may not have left a detectable epidemiological footprint on dengue. Environmental catastrophes involving flooding and land disruption can amplify arboviral transmission through several pathways: stagnant water accumulates in affected landscapes and creates new breeding sites; disruption of water supply forces households to store water in open containers; and displacement of populations may bring susceptible individuals into contact with circulating viral strains (Acosta-España et al. 2024; Rupa and Hossian 2024).

The Brumadinho tailings release disrupted riparian ecosystems along the Paraopeba River valley and affected water supply across multiple municipalities (Vergilio et al. 2020), and some of these conditions may have locally favoured *Aedes aegypti* breeding. Yet any such effect was probably diluted at the municipality level by the dominant state-wide epidemic wave of 2019. The predominantly rural, low-density character of much of the Paraopeba basin also limits the transmission amplification that typically occurs in densely urbanised settings, where vector populations and human-mosquito contact rates are substantially higher (Bohm et al. 2024).

The analytical pipeline used here - Pearson correlation-based hierarchical clustering combined with PERMANOVA ordination - has been applied in recent dengue epidemiology work across Brazil (Baes Pereira et al. 2025). The absence of significant cluster enrichment for Paraopeba basin municipalities after false discovery rate correction confirms that exposed municipalities do not form a coherent epidemiological group distinct from their regional neighbours. The significant Year:Health Region interaction (p < 0.001) in the mixed-effects model further reflects the well-known spatiotemporal heterogeneity of dengue spread in Brazil, with the intensity and timing of peaks varying across health regions from year to year. That said, the borderline contrast for basin municipalities (p = 0.088) deserves attention: it falls just short of conventional thresholds and hints at a modest local signal that a study with a larger set of exposed municipalities might be better powered to detect.

Several limitations temper these conclusions. Our analysis relies on passive surveillance data from SINAN, which is subject to well-documented underreporting - a problem that worsened during the COVID-19 pandemic, which overlapped with part of the post-event period (Oliveira Roster et al. 2024; Coelho et al. 2016). If underreporting was disproportionately higher in disaster-affected municipalities - owing to disruption of healthcare services and surveillance capacity (Vidal, De Cássia Costa Da Silva, and Zucchi 2024) - the true incidence in those areas may be underestimated, potentially masking a real local effect. The absence of entomological, climate, and serotype data further limits mechanistic interpretation. The COVID-19 pandemic introduced additional confounding by disrupting vector control activities across the state (Rotondo De Araújo et al. 2023; Borre et al. 2022), a trend that may have independently inflated post-2019 dengue incidence across the state.

Resolving the open question of whether the dam collapse had any localised effect on dengue will require studies with finer spatial and temporal resolution, individual-level exposure data, and entomological assessments collected prospectively in the aftermath of the disaster. Serotype surveillance data would help distinguish surges driven by the introduction of new DENV lineages from those reflecting increased host susceptibility.

In summary, dengue dynamics in Minas Gerais between 2014 and 2023 were shaped primarily by health-region structure and state-wide epidemic forces, with no statistically significant independent contribution from the 2019 Brumadinho dam collapse. The findings point to the value of robust comparative designs when assessing the epidemiological consequences of environmental disasters, and reflect the difficulty of isolating localised signals against a background of concurrent large-scale outbreaks.

## 5. Conclusion

This study characterized the spatiotemporal dynamics of dengue incidence across municipalities of Minas Gerais from 2014 to 2023 and evaluated the hypothesis that the 2019 Brumadinho dam collapse altered dengue transmission patterns in the Paraopeba River basin. Our main findings can be summarized as follows: (i) dengue dynamics in Minas Gerais are predominantly structured by health region, reflecting shared large-scale climatic and ecological drivers; (ii) temporal variability was governed by state-wide epidemic forces, with 2023, 2019, and 2016 emerging as the dominant epidemic years; and (iii) municipalities within the Paraopeba basin showed no statistically distinct epidemiological trajectory relative to control municipalities after the dam disaster, when health region effects and the broader temporal trend were accounted for.

These results do not exclude the possibility that the Brumadinho disaster had localized, short-term effects on vector habitats or human vulnerability to dengue that were too fine-grained to be detected at the annual, municipality-level resolution of this study. However, they do indicate that no lasting or large-scale disruption of regional dengue dynamics can be attributed to the event independently of concurrent state-wide epidemic drivers. The observed increase in dengue incidence across Minas Gerais after 2019 is better explained by national epidemic cycles, serotype dynamics, and broader socio-environmental determinants than by the localized environmental impact of the dam collapse.

From a public health perspective, our findings reinforce the importance of health region as an operational unit for dengue surveillance and response planning in Minas Gerais, and highlight the need for strengthened and adaptive surveillance systems capable of detecting localized epidemic signals against the background of large, synchronous state-wide outbreaks. Future studies using finer spatial and temporal resolution, quasi-experimental designs, and integrated environmental and entomological data will be essential to fully characterize the public health footprint of disasters on vector-borne disease dynamics in affected communities.

## Authors contribution

Conceptualization: GRF, WKO, MLPC, CRMF, CES. Data curation: GRF, WKO. Formal Analysis, Methodology and Investigation: GRF. Writing – Review and Editing: GRF, ABMV, PLCF, WKO. ERGRA, BCL, CRMF, MLPC, CES. Project Administration: ABMV, MLPC, CES. Funding Acquisition: CRMF, CES, MLPC.

## Supporting information

Strobe Checklist

## Data Availability

The data utilized in this project was sourced from SINAN and is archived at https://github.com/Infection-control/projetovale_wanderson.

## Acknowledgments

The funding for this project was provided through reparation funds originating from Vale mining company.

## Notes

### Author Declarations

The analyses in this study were conducted exclusively using aggregated/summary data. The primary data source is the Brazilian Notifiable Diseases Information System (SINAN — Sistema de Informaç~o de Agravos de Notificaç~o), which is a publicly available, government-managed surveillance database maintained by the Brazilian Ministry of Health. SINAN records are anonymised at the point of entry into the system: no personal identifiers — such as names, addresses, or identification numbers — are accessible to researchers. The variables available to users are restricted to epidemiologically relevant fields (e.g., disease code, date of symptom onset, municipality of residence, and basic demographic categories), in full compliance with Brazilian data protection legislation. For the purposes of this study, individual notifications were aggregated to the municipality-year level prior to analysis, yielding a dataset of annual dengue case counts per municipality. No individual-level data were used in, or are reported in, the manuscript. The resulting dataset therefore contains no information that could be used to identify any individual patient.

