## Supplementary material for "Dengue spatiotemporal patterns in Minas Gerais, Brazil, 2014 – 2023: regional epidemic forces dominate over the environmental impact of the Brumadinho dam collapse": Strobe Checklist

*Strengthening the Reporting of Observational Studies in Epidemiology*

**Study design: Ecological (cross-sectional spatiotemporal analysis)**

| **Item** | **STROBE Recommendation** | **Notes** | **Location in Paper** | **Status** |
| --- | --- | --- | --- | --- |
| **TITLE AND ABSTRACT** | | | | |
| **1a** | Indicate the study's design with a commonly used term in the title or abstract. | Title states "spatiotemporal analysis"; study design (ecological) identified in abstract Methods. | *Title; Abstract — Methods paragraph* | **✓ Done** |
| **1b** | Provide in the abstract an informative and balanced summary of what was done and what was found. | Structured IMRaD abstract with Background, Methods, Results, and Conclusions paragraphs. | *Abstract* | **✓ Done** |
| **INTRODUCTION** | | | | |
| **2** | Explain the scientific background and rationale for the investigation being reported. | Dengue epidemiology in Brazil, spatiotemporal approaches, Brumadinho disaster, and knowledge gap all described. | *Section 1 — Introduction (¶1–4)* | **✓ Done** |
| **3** | State specific objectives, including any prespecified hypotheses. | Final paragraph of introduction states the hypothesis explicitly: dam collapse may have influenced dengue dynamics in affected municipalities. | *Section 1 — Introduction (¶5)* | **✓ Done** |
| **METHODS** | | | | |
| **4** | Present key elements of study design early in the paper. | Ecological spatiotemporal study design stated at the opening of Section 2. | *Section 2.1 — Study design and data sources* | **✓ Done** |
| **5** | Describe the setting, locations, and relevant dates, including periods of recruitment, exposure, follow-up, and data collection. | Minas Gerais state described; study period 2014–2023; Brumadinho dam collapse date (25 Jan 2019) given. | *Section 2.1; Introduction ¶3* | **✓ Done** |
| **6** | Eligibility criteria, and the sources and methods of selection of participants. For matched studies, give matching criteria and number of exposed and unexposed. | 67 municipalities included: 26 Paraopeba basin (formally designated as affected via technical-political process tied to Vale reparation investments) and 41 regional controls (same health regions, not designated). Selection rationale and group sizes stated explicitly. Supplementary Table S1 lists all municipalities. | *Section 2.1 — Study design (¶2); Supplementary Table S1* | **✓ Done** |
| **7** | Clearly define all outcomes, exposures, predictors, potential confounders, and effect modifiers. Give diagnostic criteria, if applicable. | Dengue incidence (outcome) and Paraopeba basin status (exposure) defined. Potential confounders (urbanisation, socioeconomic factors, COVID-19) acknowledged in Discussion; not formally controlled as covariates in the main model. | *Section 2.2; Discussion — Limitations paragraph* | **⚠ Partial** |
| **8** | For each variable of interest, give sources of data and details of methods of assessment (measurement). Describe comparability of assessment methods if there is more than one group. | Outcome data from SINAN described; population denominator from IBGE 2022 census; municipality metadata described. | *Section 2.1; Section 2.2* | **✓ Done** |
| **9** | Describe any efforts to address potential sources of bias. | Passive surveillance underreporting and COVID-19 pandemic effects acknowledged as limitations. No formal sensitivity analysis performed. | *Discussion — Limitations paragraph* | **⚠ Partial** |
| **10** | Explain how the study size was arrived at. | No power calculation performed. All 67 municipalities involved with the disaster included as a census. Study period constrained by SINAN database start date (2014). Stated explicitly. | *Section 2.7 — Study size* | **✓ Done** |
| **11** | Explain how quantitative variables were handled in the analyses. If applicable, describe which groupings were chosen and why. | Z-score standardisation of incidence described; year treated as categorical factor in mixed model; cluster number determined by silhouette method. | *Section 2.2; Section 2.3; Section 2.5* | **✓ Done** |
| **12a** | Describe all statistical methods, including those used to control for confounding. | Hierarchical clustering, PERMANOVA, NMDS, PCA, linear mixed-effects model with municipality random effect all described with package citations. | *Section 2.3–2.5* | **✓ Done** |
| **12b** | Describe any methods used to examine subgroups and interactions. | Interaction terms (Year × Paraopeba basin × Health region) included in mixed-effects model; pre/post contrasts estimated within each group via emmeans. | *Section 2.5* | **✓ Done** |
| **12c** | Explain how missing data were addressed. | Municipality-years with no reported cases assigned zero; no other missing data handling described. | *Section 2.2* | **⚠ Partial** |
| **12d** | If applicable, describe analytical methods taking account of sampling strategy. | Not applicable: complete census of relevant municipalities, no sampling. | *N/A* | **✗ N/A** |
| **12e** | Describe any sensitivity analyses. | No sensitivity analyses performed. Acknowledged as a limitation. | *Discussion — Limitations; Future directions* | **⚠ Partial** |
| **RESULTS** | | | | |
| **13a** | Report numbers of individuals at each stage of study (e.g. numbers potentially eligible, examined for eligibility, confirmed eligible, included in the study, completing follow-up, and analysed). | 67 municipalities included (26 Paraopeba basin, 41 regional controls). Municipalities excluded after zero-variance filter noted in supplementary script. Total listed in Supplementary Table S1. | *Section 2.7; Supplementary Table S1; Supplementary Script S1* | **✓ Done** |
| **13b** | Give reasons for non-participation at each stage. | Not applicable: all eligible municipalities included; no refusal or dropout possible in an ecological study using registry data. | *N/A* | **✗ N/A** |
| **13c** | Consider use of a flow diagram. | Flow diagram not included. For an ecological study using complete registry data, a PRISMA-style diagram is not standard practice. | *Not included* | **⚠ Partial** |
| **14a** | Give characteristics of study participants (e.g. demographic, clinical, social) and information on exposures and potential confounders. | Municipality characteristics (health region, study group, population, annual case counts) provided in Supplementary Table S1. | *Supplementary Table S1* | **✓ Done** |
| **14b** | Indicate number of participants with missing data for each variable of interest. | No individual-level data; no missing municipality records. Municipalities with zero recorded cases in a given year were retained with count = 0. | *Section 2.2* | **✓ Done** |
| **15** | Report numbers of outcome events or summary measures. | Epidemic peaks (2015–2016, 2019, 2023) described; case counts in Supplementary Table S1; forest plot (Supp. Fig. S2) compares pre/post counts. | *Section 3.1; Supplementary Table S1; Supplementary Fig. S2* | **✓ Done** |
| **16a** | Give unadjusted estimates and, if applicable, confounder-adjusted estimates and their precision (e.g. 95% confidence interval). Make clear which confounders were adjusted for and why they were included. | PERMANOVA R² and p-values; mixed-model pre/post contrasts with estimates, SE, df, t-ratio, and p-values reported in Tables 2 and 3. Health region included as a covariate. | *Tables 2–3; Section 3.3–3.4* | **✓ Done** |
| **16b** | Report category boundaries when continuous variables were categorised. | Year treated as a categorical factor (one level per year, 2014–2023). No continuous-to-categorical conversion of other variables. | *Section 2.5* | **✓ Done** |
| **16c** | If relevant, consider translating estimates of relative risk into absolute risk for a meaningful time period. | Odds ratios from forest plot (Supp. Fig. S2) and Z-score contrasts reported. Absolute incidence rates provided in Supplementary Table S1. | *Supplementary Fig. S2; Supplementary Table S1* | **✓ Done** |
| **17** | Report other analyses done — e.g. analyses of subgroups and interactions, and sensitivity analyses. | Year × Health region interaction significant (Table 2). No sensitivity analyses performed; acknowledged as a limitation. | *Table 2; Discussion — Limitations* | **⚠ Partial** |
| **DISCUSSION** | | | | |
| **18** | Summarise key results with reference to study objectives. | Discussion opens with a clear summary of the main finding relative to the stated hypothesis. | *Discussion ¶1* | **✓ Done** |
| **19** | Discuss limitations of the study, taking into account sources of potential bias or imprecision. Discuss both direction and magnitude of any potential bias. | Underreporting, COVID-19 confounding, differential surveillance disruption, ecological design limitations, and absence of entomological/serotype data all discussed. | *Discussion — Limitations paragraph* | **✓ Done** |
| **20** | Give a cautious overall interpretation of results considering objectives, limitations, multiplicity of analyses, and results from similar studies. | Null finding interpreted cautiously; borderline p = 0.088 noted as possible underpowered local signal; results contextualised against Machado et al. (2025). | *Discussion ¶3–5; Conclusion* | **✓ Done** |
| **21** | Discuss the generalisability (external validity) of the study results. | Findings are specific to Minas Gerais. Transferability to other disaster-affected regions not explicitly discussed. | *Discussion — Future directions* | **⚠ Partial** |
| **OTHER INFORMATION** | | | | |
| **22** | Give the source of funding and the role of the funders for the present study and, if applicable, for the original study on which the present article is based. | Funding statement added: reparation funds from Vale mining company. | *Acknowledgments* | **✓ Done** |
| **Legend: ✓ Done** ⚠ Partial / requires attention ✗ Not applicable | | | | |

*Checklist based on: Vandenbroucke JP et al. Strengthening the Reporting of Observational Studies in Epidemiology (STROBE): explanation and elaboration. PLoS Med. 2007;4(10):e297. doi:10.1371/journal.pmed.0040297*
